# Phenotyping Adolescent Endometriosis: Characterizing Symptom Heterogeneity Through Note- and Patient-Level Clustering

**DOI:** 10.1101/2025.02.10.25321215

**Authors:** Rebecca M Cohen, Emily Leventhal, Nivedita Nukavarapu, Victoria Lazarov, Sarriyah Hanif, Michal A Elovitz, Kimberly B Glazer, Ipek Ensari

## Abstract

**Introduction:** Pelvic pain (dysmenorrhea and non-menstrual) is the most common presentation of adolescent endometriosis, but symptoms vary between and within patients. Other presentations, such as gastrointestinal (GI) symptoms, are often misattributed, leading to diagnostic delays. Patients incur frequent primary and specialty care visits, generating multiple and diverse clinical notes. These offer insights into disease trajectory and symptom heterogeneity, which can be rigorously investigated using clustering methods. This study aims to 1) evaluate phenotypes using electronic health records (EHRs) and 2) compare two clustering models (note-vs patient-level) for their ability to identify symptom patterns.

**Methods:** We queried the Mount Sinai Data Warehouse for clinical notes from patients aged 13-19 years with a SNOMED endometriosis diagnosis, yielding an initial sample of 7,221 notes. A randomly selected subsample was annotated with 12 disease-relevant labels, including symptoms, hormone use, and medications. The final analytic sample included 695 notes from 26 unique patients. Pelvic pain, dysmenorrhea, chronic pain, and GI symptoms were selected as model predictors based on principal component analysis. Two unsupervised machine learning (ML) methods were then applied for note-vs patient-level analyses: Partitioning Around Medoid (PAM) and Multivariate Mixture Models (MGM).

**Results:** The PAM model identified K=3 clusters with average silhouette width of 0.76, indicating strong between-cluster separation. The “feature-absent” (abs) phenotype (76%) was distinct for absence of all 4 features. The “classic” phenotype (8%) exhibited pelvic pain, dysmenorrhea, and chronic pain. The “GI” phenotype (16%) was dominated by GI symptoms. The MGM identified K=2 stable patient-level clusters (Δ weighted model deviance = -224.93 from K=2 to 3) with a mean cluster membership probability of 0.97: A “classic” phenotype (50%), characterized by pelvic pain and chronic pain, and a “non-classic” phenotype (50%), defined by the absence of these features. PAM-based classic phenotype had significantly higher rates of hormonal intervention (78% vs 26% abs, 49% GI) and pain medication (68% vs 9% abs, 14% GI). For the patient-level, the classic phenotype also had higher average rates per person of hormonal therapy (26% vs 7%) and prescription pain medications (27% % vs 9%) (p<0.01 for all).

**Conclusions:** Both methods captured classic and non-classic phenotypes, with the note-level model uniquely identifying a feature-absent group. The classic phenotype’s link to higher hormonal and pain intervention underscores the importance of recognizing non-classic symptoms. This study, the first to directly compare note-and patient-level clustering of EHR notes in endometriosis, demonstrates the ability to detect the less clinically recognizable phenotypes. This proof-of-concept can be applied to larger datasets to refine phenotype identification, aiding in earlier diagnosis.

## Introduction

Endometriosis is a chronic gynecological condition characterized by the presence of endometrial-like tissue outside the uterus, affecting approximately 10% of reproductive-age women worldwide^1^. This prevalent disease imposes a significant burden on patients, healthcare systems, and society at large. Women with endometriosis often experience debilitating symptoms such as chronic pelvic pain, dysmenorrhea, and infertility, leading to reduced quality of life and substantial economic costs due to healthcare utilization and lost productivity^2^. Despite its prevalence and impact, endometriosis remains underdiagnosed and undertreated, with an average delay of 7-10 years from symptom onset to definitive diagnosis^1, 3^.

Up to two-thirds of patients with endometriosis report onset of symptoms during adolescence^1, 4^. It is the leading cause of dysmenorrhea and chronic pain in adolescents, significantly impairing both physical and psychosocial functioning^15^. Adolescent endometriosis presents unique diagnostic and management challenges, as symptoms may be atypical or dismissed as normal menstrual discomfort^3^. Pelvic pain is the most common presenting symptom in adolescents (present in 63% of adolescents), presenting as both dysmenorrhea and/or non-menstrual pelvic pain. Gastrointestinal (GI) and bowel symptoms are less recognized but not uncommon presenting symptoms. Overall, the heterogeneity in symptoms between- and within patients over time, and the invasive nature of diagnostic laparoscopy, contribute to the delays in diagnosis. Early identification of specific symptom patterns (phenotypes) could enable more timely interventions, and thus greatly improve quality of life in this patient population.

Given the complex and heterogeneous nature of endometriosis, particularly in adolescents, exploratory data analysis techniques such as clustering can be valuable tools for uncovering patterns and subgroups within the patient population. Clustering algorithms can identify groups of patients with similar symptom profiles, comorbidities, or treatment responses, potentially leading to more targeted and personalized approaches to diagnosis and management^6^. Clustering has been successfully used in other medical fields such as asthma^7, 8^, obesity^9^, chronic pain syndromes^10, 11^ and sleep disorders^12^. Additionally, clustering analysis may reveal previously unrecognized associations between symptoms or clinical features, generating new hypotheses about disease mechanisms or progression in young patients. In particular, application of these techniques to data extracted from electronic health records (EHRs) can facilitate identification of distinct phenotypes or risk profiles that could inform clinical decision-making and guide future research efforts.

The widespread adoption of EHRs presents a valuable opportunity to better characterize and elucidate adolescent endometriosis using real-world clinical data on demographics, symptoms, diagnostic tests, treatments, and outcomes^13^. Importantly, the unstructured clinical notes often contain rich descriptions of symptoms and clinical findings that may not be adequately represented in structured data fields. This is particularly relevant for endometriosis, which tends to be under-documented within the EHRs^14^. Leveraging these unstructured data through natural language processing (NLP) techniques can provide deeper insights into the diverse symptom patterns and clinical presentations of adolescent endometriosis^13^. Furthermore, the longitudinal nature of EHR data allows for the study of disease progression and treatment outcomes over time, potentially identifying factors associated with earlier diagnosis or better management strategies.

Clustering of clinical notes introduces important considerations. First is the decision of whether clinical notes should be treated independently (note-level analysis) or grouped by patient (patient-level analysis). For example, the commonly used K-means algorithm partitions observations (i.e., individual notes) into groups, isolating the note from patients’ clinical history. These notes can represent a “snapshot” of a patient’s disease at a given moment – such as a brief doctor’s visit where the patient’s medical history is not a primary focus or when previous medical history for the patient is not available for review. In contrast, hierarchical clustering algorithms partition the data at the patient level, capturing symptom patterns aggregated across all notes belonging to an individual. This could be closer to a clinical scenario where the physician can track a patient’s ongoing care over time. Accordingly, both clustering approaches have clinical relevance and potential use from an application perspective, depending on the use case scenario.

To date, no study has compared note-level and patient-level clustering methods in any medical field toward this endeavor. This study aims to address this gap through a comparative unsupervised learning framework to identify potential subtypes (“phenotypes”) of adolescent endometriosis by leveraging EHR data. Our objectives are to: 1) Implement unsupervised machine learning (ML) to identify adolescent endometriosis subtypes (phenotypes), and 2) Compare phenotypes generated from note-level clustering (treating each note as a separate entity) versus patient-level clustering (aggregating symptom patterns across all notes for each patient).

## 1. Methods

### 1.1. Study sample

Clinical notes from patients aged 13-19 with a SNOMED diagnosis of endometriosis were obtained from the Mount Sinai Data Warehouse. The query included all clinical notes per patient except for telephone encounters, resulting in 7,221 notes from 122 patients. For this proof-of-concept study, a random sample of 700 notes were selected for annotation and labeling. Exclusion criteria included having fewer than 3 clinical notes, due to clustering model convergence restraints, yielding an analytic sample of 695 notes from 26 unique patients. All study procedures were approved by the Institutional Review Board at Icahn School of Medicine at Mount Sinai (ISMMS) (IRB= STUDY-22-01581-MOD0010).

### 1.2. Definitions and outcomes

Each note was manually annotated by domain experts in isolation from the other notes from the same patient (i.e. at the “note level”) with 12 disease relevant labels including symptoms, hormone use, and medications: endometriosis, pelvic pain, pelvic tenderness, abdominal pain, abdominal tenderness, gastrointestinal symptoms, dysmenorrhea, dyspareunia, pain severity, pain chronicity, pain medication use, and therapy utilization (includes hormonal therapy, IUD, and/or birth control). These labels were chosen based on the literature on endometriosis definitions and with guidance from OBGYN domain experts (SH, VL). Annotations were made according to any reference to current or prior experience of the patients within the individual note, according to the following study specific definitions. *Endometriosis* was defined as a mention of endometriosis confirmed with Laparoscopy within the individual clinical note. *Pelvic pain* was defined as any non-cyclical, non-menstrual pelvic pain or discomfort, while *dysmenorrhea* was defined as cyclic cramping, pain or discomfort. *Abdominal pain* was annotated as any non-cyclic abdominal pain superior to the pelvis. *Abdominal tenderness* and *pelvic tenderness* both refer to pain or discomfort elicited on clinical physical exam. *Gastrointestinal symptoms* include all bowel symptoms (diarrhea, constipation, gas, irritable bowel syndrome), as well as irritable bowel disease (IBD), nausea and gastritis. *Chronic pain* was defined as a mention of pain or discomfort lasting 3 months or longer, consistent with pelvic pain studies^15^. *Pain severity* was defined as either a mention to ‘severe pain’ or a pain rating equal to or greater than 7/10 on the numeric pain rating scale, or pain that is described as debilitating to everyday activities. *Pain medication use* was specified as a note-specific reference to current, past, or future pain medication, and was further stratified into over-the-counter pain medications versus prescription pain medications. Similarly, *hormonal, IUD, and birth control* was stratified into respective categories, annotated if mentioned in the isolated note. All variables were eventually dichotomized into binomial variables.

### 1.3. Clustering and data analysis

With the overall goal of identifying the most clinically relevant independent predictors that inform clustering behavior with the greatest ability to discriminate between phenotypes based on symptoms, we utilized previously described methods and adapted for our data^12, 16-18^. Briefly, this involves dimension reduction and variable selection via multiple correspondence analysis (MCA), which identifies predictors that simultaneously explain the largest amount of variability in the data that have the least amount of correlation. Then, iterative model fitting is used to identify the best-fitting model based on the model fit indices appropriate for the model type being used (i.e., point variability, silhouette width and within-cluster sum of squares; WCSS, Bayesian fit indices).

#### 1.3.1. MCA and variable selection

We conducted an MCA to explore association patterns among the binomial variables and reduce dimensionality. MCA is similar to the more commonly used principal component analysis. It was selected for this analysis as it is better suited for the binary nature of our data^19-21^, allowing us to capture relevant dimensions where data variability is maximized while addressing potential collinearity.

The number of dimensions to retain was determined based on the widely accepted criterion that retained dimensions should account for more than 70% of the cumulative variance^21, 22^. We then inspected each variable’s contribution (%), coordinate (i.e., correlation) values, and cosine squared (cos^2^) values (i.e., quality of representation) within the selected dimensions to identify key predictors.

The most informative variables from the selected MCA dimensions were chosen for clustering analysis to estimate phenotypic patterns. In instances where two predictors showed similar associations with a dimension but were highly collinear, we retained the predictor that improved cluster distinction and model feasibility.

### 1.4. Note-level clustering analyses

For note-level clustering, we used the partitioning around medoids (PAM)^23^ algorithm, which is the recommended method for categorical and binary variables^24, 25^. This involves first standardizing the data by converting to a matrix of average of partial dissimilarities between pairs of data points^26^ based on root-sum-of-squares (Euclidean) distances. The PAM algorithm uses these similarities/dissimilarities from the distance matrix to identify note clusters based on the selected features. This method identifies medoids (representative data points within each cluster), unlike K-Means which uses centroids—the mean of all data points in a cluster—making it better suited for binary data^25, 27^. Furthermore, it is more robust against noise and outliers,^28, 29^ therefore making it better suited for smaller datasets^24, 29, 30^. We used the “cluster” library in R statistical software for the analyses^31^.

#### 1.4.1. Model Selection and Fit Assessment

The overall model fit was assessed based on the model fit indices of cluster cohesion, separation, and consistency across subsets of data. Silhouette width values range from -1 to 1 and provide a measure of how well each note aligned within its assigned cluster, with values closest to 1 indicating better alignment. This value balances within-cluster cohesion and separation from other clusters and can be utilized to select the ‘appropriate’ number of clusters.^24, 32^ WCSS quantifies cluster compactness by calculating the distance of notes to their cluster’s medoid, with lower values indicating tighter consistency within clusters^33^. Lastly, stability analysis by bootstrapping methods (measured by Jaccard indices; JI) was conducted to verify that clustering results were reproducible across random subsets, confirming that the identified clusters represented robust symptom patterns^34^. JI values range from 0 to 1, with values closest to 1 indicting higher stability.

### 1.5. Patient-level clustering analyses

For the patient-level clustering, we applied an unsupervised ML method based on multivariate mixture generalized linear mixed models (MMGLMMs)^35, 36^. This approach was chosen for its ability to accommodate data that span multiple variables and recognize co-occurrence patterns over multiple notes. It specifically models each patient’s data through a set of latent random effects (i.e. predictors), estimating the likelihood of cluster membership (referred to as individual component probability; ICP), using Markov Chain Monte Carlo (MCMC)-based Bayesian inference. The approach involves defining a prior distribution for model parameters and subsequently inferring cluster membership by analyzing the posterior sample distribution generated from the MCMC. The posterior median probabilities are used to calculate each patient’s probability of belonging to a specific cluster. For our model, we included the number of notes as a covariate, 6,000 MCMC samples and discarded the first 1,000 as burn in to enable stability in chain convergence^37^.

The MMGLMM is an effective technique for patient-level clustering analysis for several reasons. First, the approach is well-suited to handle data types that are sampled irregularly across multiple time points and accounts for correlations among repeated measures from the same individual by incorporating random effects^12, 36^. This model does not rely on the assumption of classical normality; instead, it allows for different normal mixtures in the random effects distribution for each cluster, enhancing model fit and providing robustness against potential misspecification of random effects^12^. The analysis was performed using the “MixAK” library in R^35^, and further details on MMGLMM methodology are described elsewhere^35, 36^.

#### 1.5.1. Assessment of model fit

Overall model fit was assessed using the penalty of expected deviance (PED) index and the posterior distribution of model deviances (D), as recommended for MMGLMMs ^35, 38^. The PED combines the expected model deviance with the model complexity penalty term *optimism, p(opt*)^39^. Lower PED values indicate better model fit. By comparing PED values across multiple MCMC-estimated models, it can be used to inform model choice based on minimum loss. The metric has been applied across various sample sizes and parameter settings in similar phenotyping studies across healthcare contexts^12, 39^. As an additional approach to guide model selection, we also examined deviance distributions to compare models, following established recommendations.

#### 1.5.2. Model convergence diagnostics

The effective sample size (ESS) and split-Ř statistic were used to assess the convergence of the MCMC simulations, consistent with published guidelines.^40, 41^ The ESS measures whether the number of MCMC iterations is sufficient to provide stable estimates, with ESS > 100 indicating adequate estimator accuracy^41^. On the other hand, the split-Ř statistic evaluates the mixing of the Markov chains with a score of 1.1 or less indicating desirable convergence^40^.

### 1.6. Evaluation of estimated phenotype profiles for note- and patient-level

For both note-level and patient-level analyses, post-hoc Chi-squared tests were performed to both confirm inter-cluster differentiation in the predictor variables and examine phenotype-level differences in clinical features not included in the clustering algorithms. P values less than 0.05 were considered significant.

### 1.7. Comparison of note- and patient-level clusters

We compared the model-identified clusters, at the individual- and cluster levels. Specifically, we evaluated the characteristics of the clusters from both models for alignment across the 2 approaches, i.e., number and size of the clusters, number of notes and patients in each, similarities and differences in feature contribution to each cluster. We then inspected each patient’s cluster assignment for alignment across the 2 approaches. We further compared cluster alignment using visualization methods to explore the note-level cluster spread (number of note-level clusters and frequencies of notes) for each individual. Finally, we computed the frequency of each feature per cluster and patient to further characterize the clusters identified from each model.

## 2. Results

### 2.1. Sample characteristics

Sample characteristics are provided in **Table 1**. The analytic sample includes 695 notes from 26 adolescent patients with a SNOMED diagnosis of endometriosis. Patients individually contribute an average of 26.7 +/- 41.3 with a large range of 3-214, capturing a wide range of clinical encounters. An average of 3.8 (SD 2.8, Range 1-12) departments are represented per patient. The average patient age at the time of the first note is 17.3 +/- 2.3) and 20.1 +/- 2.8 at the last note. 44% of the patients self-identified as white, 19% as black or African American, and 38% as other.

**Table 1.** Study participant characteristics (N =26)

| Characteristic | Average +/- SD | Minimum | Maximum |
| --- | --- | --- | --- |
| Age |  |  |  |
| At first note | 17.3 +/- 2.3 | 10.7 | 20.5 |
| At last note | 20.1 +/- 2.8 | 15.9 | 26.1 |
| Substance Use | 0 | **0** | 0 |
| # Notes per patient | 26.7 +/- 41.3 | **3** | 214 |
| # Departments per patient | 3.8 +/- 2.8 | 1 | 12 |
| Race | Count | Proportion |  |
| White | 11 | 0.42 |  |
| Black or African American | 5 | 0.19 |  |
| Other | 10 | 0.38 |  |

Frequencies of the 12 manually annotated features were calculated at both the note- and patient levels, reported in **Table 2**. Notably, GI symptoms were the most frequently reported outcome (appearing in 125 notes with an average of 4.81 per person), followed by abdominal pain (120 total notes, avg. 4.62 per person), and then pelvic pain (94 total, avg. 3.62 per person). On the other hand, dyspareunia was the least frequently documented symptom (11 total mentions with an average of 0.42 per person), followed by pelvic tenderness (33 total mentions, avg 1.27 mentions/person) and then abdominal tenderness (35 total mentions, avg 1.35 mentions/person). Regarding therapy, 102 total notes mentioned any pain medications (49 over the counter, 77 prescription) and 233 mentioned additional therapy (128 oral contraceptive (OCP), 73 IUD, and 68 hormonal therapy).

**Table 2.**
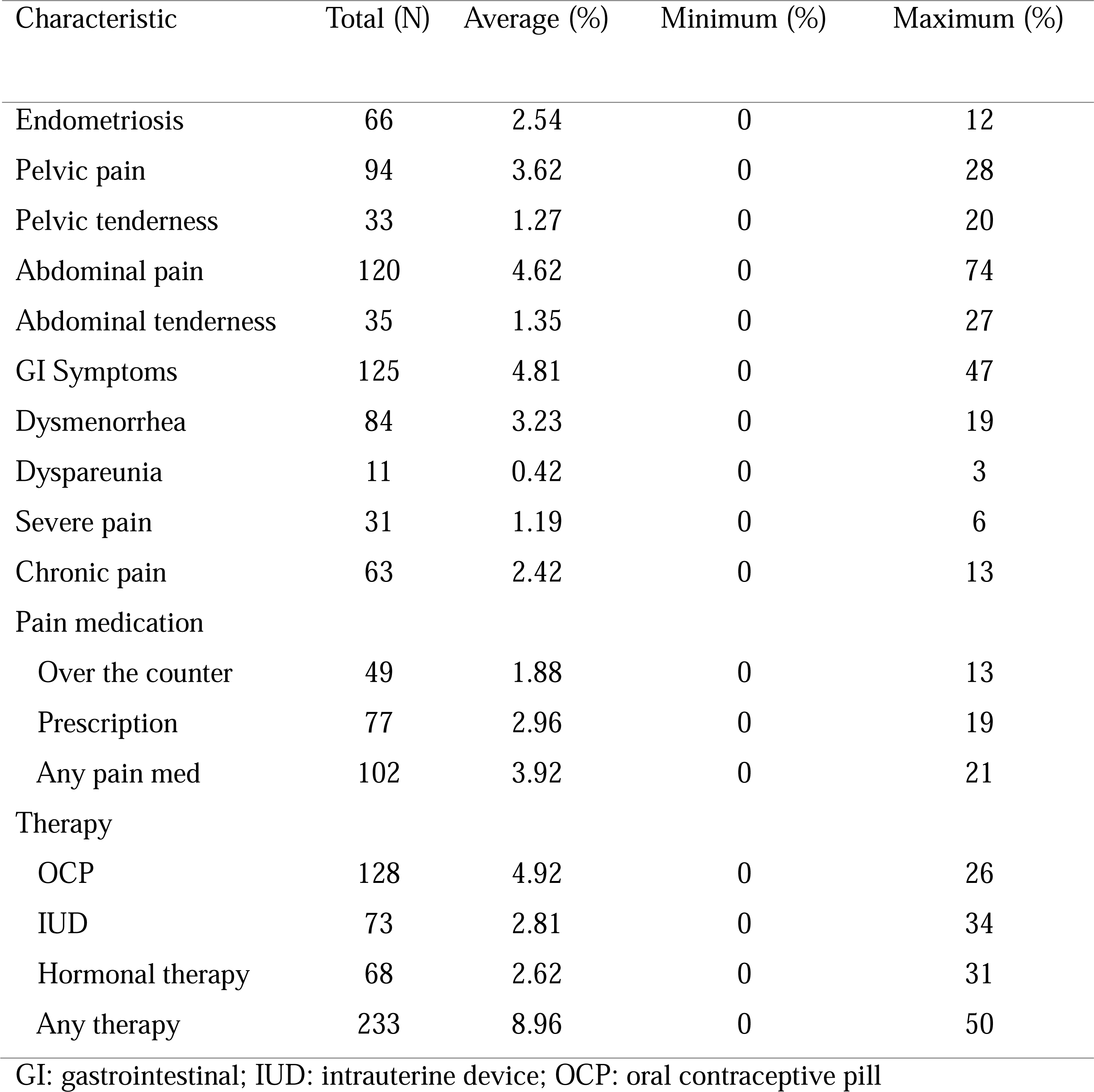
Total frequencies of each annotated feature and averages of per person frequency (Total notes = 695).

### 2.2. Feature selection and variable reduction

As shown in **Supplementary Table 1**, three symptom features are mentioned in a significantly smaller frequences when compared to the other features: dyspareunia was mentioned in 1.94% of total feature mentions, pelvic tenderness in 5.81% and abdominal tenderness in 6.16%, while the remaining symptoms appeared in 11.62% - 22.01%. Thus, these three features were not included into the MCA.

MCA results indicated that the first four dimensions individually explain more than 10.64% of the variance (36.27%, 18.56%, 13.46%, and 10.64%, respectively) and collectively account for 78.93% of the total variance, surpassing the >70% threshold for meaningful dimensionality reduction. Results detailing each symptom feature’s total contribution, representation quality, and individual coordinates (correlations) within each dimension are provided in **Supplemental Table 2.** Based on coordinate values, chronic pain (2.59) has the strongest positive association with dimension 1. Abdominal pain (1.59) and GI symptoms (1.57) are most strongly associated with dimension 2, while severe pain (3.32) has the highest correlation with dimension 3 and pelvic pain (0.87) with dimension 4. Contributions reveal that the presence of severe pain (73.66%), dysmenorrhea (77.95%), and GI symptoms (45.96%) substantially impacts the explained variance in the dimensions. Cosine squared values, representing the quality of representation, suggest that the variance for dysmenorrhea (0.95) is best captured by the associated dimensions. These results are further illustrated in **Supplementary** Figure 1.

Correlations between the highest contributing features are provided in **Supplementary Table 3**, showing high collinearity among several symptom pairs: chronic pain and pelvic pain (0.56, p<0.0001), GI symptoms and abdominal pain (0.49, p<0.0001), dysmenorrhea and chronic pain (0.44, p<0.0001) and chronic pain and endometriosis (0.44 p<0.0001).

**Dysmenorrhea**, **GI symptoms**, **pelvic pain**, and **chronic pain** are included in both models based on the evaluation of PAM model metrics and the correlations described above, combined with the goal of preserving clinically important information.

### 2.3. Note-level clustering analysis

#### 2.3.1. PAM model evaluation

The PAM clustering analysis identifies a three-cluster (K=3) solution as the optimal model for the data based on the silhouette width, WCSS, and the Jaccard index (**Supplementary Table 4**). The silhouette width improves significantly from K=2 (0.67) to K=3 (0.76, Δ(K2 to 3) =0.09) but shows marginal improvement with K=4 (0.79, Δ0.03), indicating optimal fit at K=3.^24, 31^ Similarly, the WCSS, which quantifies the compactness of clusters, decreases from 0.27 at K=2 to 0.19 at K=3 (Δ0.08), and further decreases to 0.15 at K=4 (Δ0.04), indicating tighter clustering. Calculating the Point variability and JI introduces an error at K=4, indicating model saturation at K=3.

**Supplementary Table 5** describes detailed metrics for the selected K=3 model. Notably, the silhouette width was highest for the feature-absent phenotype (0.84), indicating strong separation, while the GI phenotype and classic phenotype had moderate silhouette widths of 0.51 and 0.39, respectively. Cluster stability, assessed via the JI, demonstrated highest reproducibility for the feature-absent phenotype (0.95) but lower reproducibility for the classic and GI phenotypes.

#### 2.3.2. Note-level phenotypes

Three note-level phenotypes are displayed in **Figure 1**. Post-hoc *X*^2^ tests between phenotypes for all annotated features (including the four model predictors) are summarized in **Supplementary Table 6.** The *X*^2^ statistic from the test of independence indicated statistically significant differences (*p*<0.05) across phenotypes for all features except hormonal therapy alone, indicating strong distinctions among the identified phenotypes.

**Figure 1.**
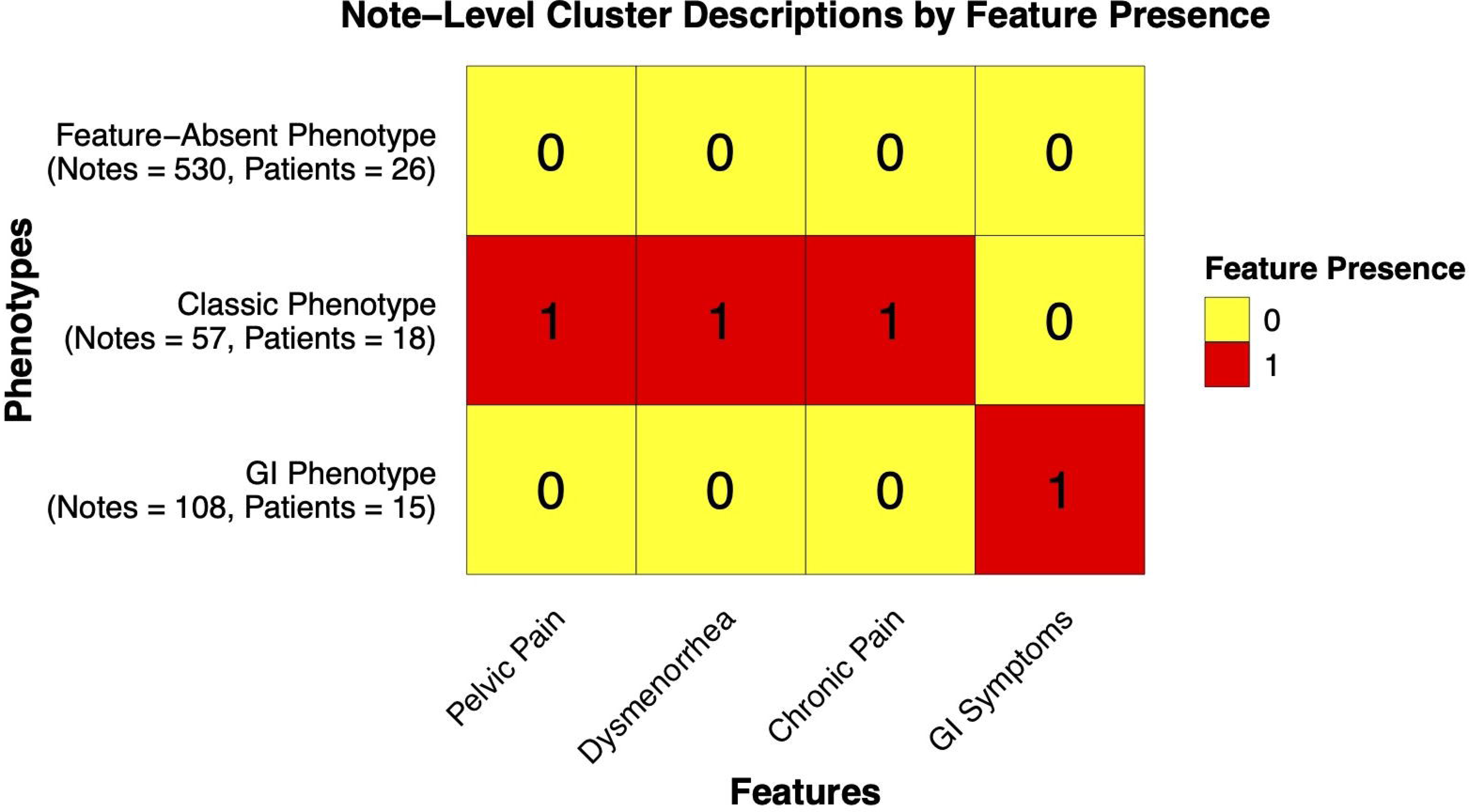
PAM-identified note-level phenotype characteristics based on the indicator variables. Abbreviations: GI: gastrointestinal

Phenotype 1, referred to as the “**feature-absent” phenotype,** constitutes the largest group, with 530 notes (76% of the sample) and includes notes from all 26 patients in the dataset. This phenotype is characterized by the absence of pelvic pain, dysmenorrhea, chronic pain, and gastrointestinal symptoms (all four model indicator variables) (**Figure 1**). Of the notes in this phenotype, the four indicator variables are mentioned at very low frequencies: only 5.3% of the notes mention pelvic pain, 4.9% mention dysmenorrhea, 0.4% mention chronic pain and 0.0% mentioned GI symptoms (p<0.001 for all) (**Supplementary Table 6**). This phenotype was further associated with lower likelihood of intervention, with only 25.9% of notes documenting a therapy (composite of OCPs and/or IUDs and/or hormonal therapies) and 9.1% reporting a pain medication (over the counter or prescription) (**Supplementary Table 6**).

Phenotype 2 (the **“classic” phenotype**) represents 8% of the notes (n=57) and includes 18 unique patients, and is defined by the presence of pelvic pain, dysmenorrhea, and chronic pain, exhibiting the hallmark symptoms of endometriosis (**Figure 1**). Regarding the frequencies of the four indicator variables in this group, chronic pain has the highest frequency (93.0%), followed by pelvic pain (89.5%), and then dysmenorrhea (70.2%). As expected, GI symptoms are documented at significantly lower frequency (29.8%). This group has the highest rates of clinical intervention, with 68.4% of notes reporting some form of pain medication, and 77.2% reporting an endometriosis therapy (OCPs and/or IUDs and/or hormonal therapies) (**Supplementary Table 6**).

Phenotype 3 (the “**GI” phenotype**) includes 108 notes (16%) from 15 patients, and is defined by the presence of GI symptoms (**Figure 1**). Of the notes in this phenotype, 100.0% mention GI symptoms, while only 13.9%, 16.7%, and 7.4% mention pelvic pain, dysmenorrhea and chronic pain, respectively. This group has lower frequencies of interventions compared to classic phenotype: 13.9% of notes document pain medications and 49.2% of notes document OCP/IUD/hormonal therapy interventions (**Supplementary Table 6**).

#### 2.3.3. Note-level phenotype associations

The associations of non-indicator clinical features with each phenotype reveal additional distinctions between the three phenotypes in terms of symptomatology and intervention patterns. Pain medication usage, both prescription and over the counter (OTC), shows distinct patterns across phenotypes in Chi-squared analysis. As described, the classic phenotype has the highest rates and the feature-absent the lowest. Post-hoc X^2^ analysis (**Supplementary Table 6**) indicates that the pain medication feature was least likely to be present in the GI phenotype, compared to the other two phenotypes (X^2^ residual = -4.43, p=0.001).

Additional therapies (including hormonal therapy, OCPs, and IUDs) are also most frequently in the classic phenotype according to Chi-squared analysis (p<0.001). Post-hoc results (**Supplementary Table 6**) shows significantly higher residuals in the feature-absent phenotype (residual 8.00, p<0.001), followed by the classic phenotype (residual -3.82, p=0.009), and lastly the GI phenotype (residual -4.36, p=0.001).

### 2.4. Patient-level clustering analysis

#### 2.4.1. MMGLMM model evaluation

A two-cluster solution provided the best fit for the patient-level data based on the PED, weighted PED (wPED), and convergence diagnostics (wPED of 1993.8 K=2 vs 2218.7 for K=3). The two-cluster model also had a lower penalty for model complexity (P(opt) = 61.1 vs. 193.8), favoring a simpler structure.

Overall cluster membership probability indicated high confidence of member assignment based on based the mean ICP of 0.97 (SD = 0.08), a median of 1.00 (mad = 0.00), and a range from 0.69 to 1.00, indicating more than half of the participants were assigned into their respective cluster with 100% certainty.

Convergence diagnostics further supported the two-cluster model, which achieves good convergence with a Gelman-Rubin convergence diagnostic of 1.0, while the three-cluster model shows instability (Gelman-Rubin convergence diagnostic= 5.32, upper CI = 13.1), suggesting that the three-cluster model did not converge.

#### 2.4.2. Patient-level phenotype characterization

Figure 2 shows the two patient-level phenotypes identified by the MMGLMM model, along with the posterior mean estimates of feature-level parameters for each phenotype. We refer to these two phenotypes as the **“classic” phenotype** and the **“non-classic” phenotype** for ease of comparison and reference throughout the sections that follow.

**Figure 2.**
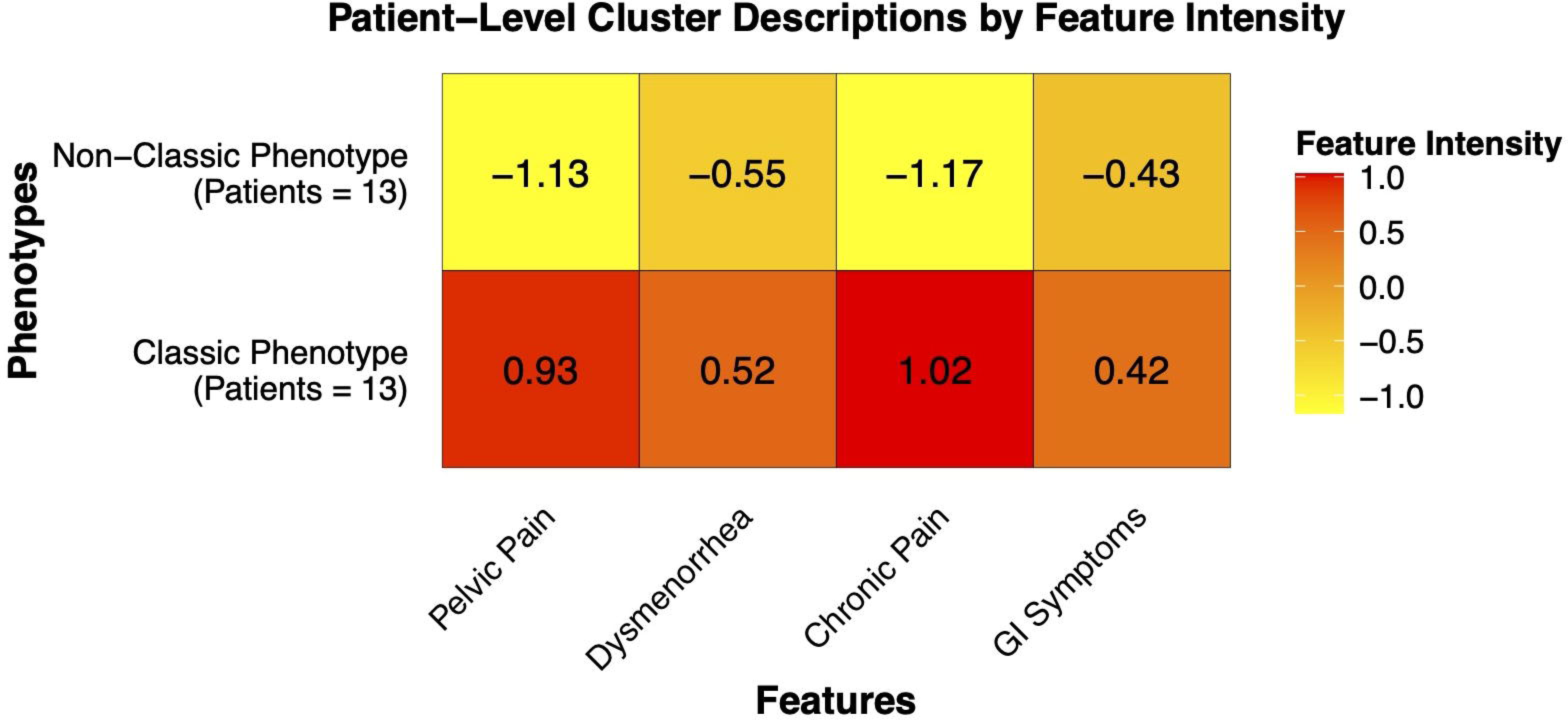
MMGLMM-identified patient-level phenotype descriptions based on indicator variable intensity (Bayesian point estimates). Each value represents the average intensity (or strength) of a specific feature for a given cluster, based on the posterior distribution after fitting the model. **Abbreviations:** GI: gastrointestinal

The classic phenotype (N=13, 50%) is characterized by higher prevalence of pain-related features, with the highest posterior mean contributions from chronic pain (1.02) and pelvic pain (0.93). Dysmenorrhea is moderately present, but it is not a defining characteristic of this phenotype based on the posterior mean estimate (0.52). GI symptoms contribute the least to the classic phenotype, with the lowest posterior mean estimate among the four features (0.42) (Figure 2).

In contrast, the non-classic phenotype (N=13, 50%) is most notably characterized by the absence of traditionally considered pain-related symptoms, as the lowest posterior mean estimates are observed for chronic pain (-1.17) and pelvic pain (-1.13). Of the four model indicator variables within this phenotype, GI symptoms have the highest posterior mean estimate in this (-0.43), indicating a relatively higher presence of GI symptoms compared to the other features (Figure 2).

Independent t-tests comparing the mean proportions of the indicator variables (**Supplementary Table 7**) confirm that the two phenotypes are most clearly distinguished by pelvic pain and chronic pain and that within the non-classic phenotype, GI symptoms are most likely to have occurrence. Compared to non-classic, the classic phenotype is associated with statistically higher mean proportions for pelvic pain (0.33 ± 0.12 vs. 0.03 ± 0.04; t = 8.4, p<0.001) and chronic pain (0.24 ± 0.17 vs. 0.03 ± 0.05; t = 4.4, p = 0.001). Finally, dysmenorrhea and GI symptoms are equally associated both phenotypes (p < 0.05 for both).

#### 2.4.3. Patient-level phenotype associations

T-tests comparing the remaining annotated features between the two patient-level phenotypes are presented in **Supplementary Table 7**. Consistent with the PAM model results, the classic phenotype shows a higher likelihood of overall therapy utilization, including hormonal therapy, OCPs, and IUDs, compared to the non-classic Phenotype (0.08 ± 0.11 vs. 0.00 ± 0.01; *p* = 0.035). Among individual therapies, only hormonal therapy differs significantly, with a higher mean proportion in the classic phenotype (0.26 ± 0.23) compared to the non-classic phenotype (0.07 ± 0.10; *p* = 0.015).

In contrast to the PAM model, overall pain medication use does not differ significantly between the two groups. However, patients in the classic phenotype are significantly more likely to use prescription pain medications (0.27 ± 0.25) compared to those in the non-classic Phenotype (0.09 ± 0.12; *p* = 0.034). Within the non-classic phenotype, the GI symptoms were associated with the highest likelihood of occurrence among all the symptom features investigated (i.e., *X^2^* statistic; See **Supplementary Table 7**).

### 2.5. Note- vs. patient-level comparisons

Figure 3 directly compares the note-level and patient-level assignments by illustrating the distribution of note-level phenotype across individual patients, stratified by patient-level phenotype. The feature-absent notes constituted the majority of notes in both patient-level phenotypes.

**Figure 3.**
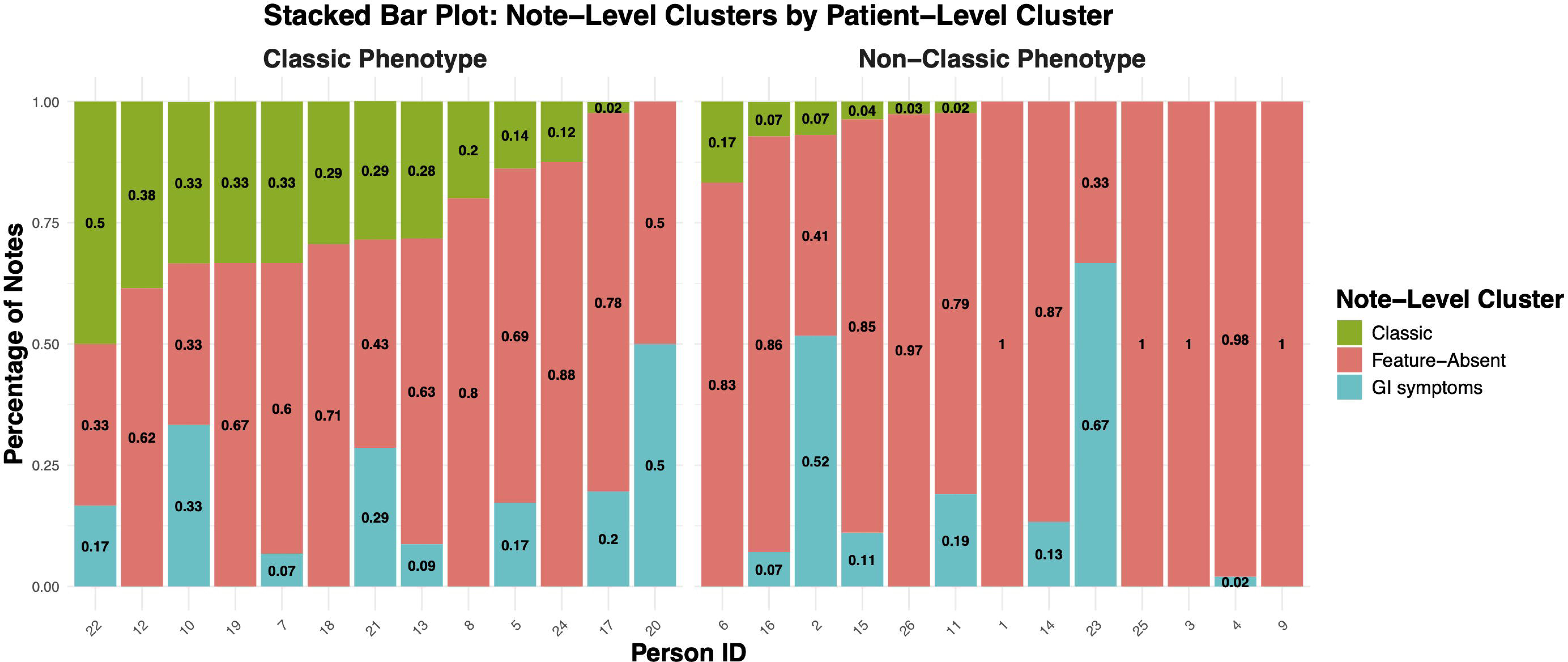
Distribution of notes for each patient along with their assignment according to the note-level (PAM) and patient-level (MMGLMM) clustering models. This figure shows the percentage distribution of notes across note-level clusters for each patient, grouped by their patient-level phenotype. Each bar represents an individual patient, with colored segments indicating the proportion of notes in each note-level cluster. The two facets highlight differences in note-level cluster distribution between the patient-level phenotypes. **Abbreviations:** GI: gastrointestinal, MMGLMM: multivariate mixture generalized linear mixed models, PAM: partitioning around medoid

The non-classic patient-level phenotype appears to capture two distinct ‘types’ of patients with few exceptions. First, it contains the four patients with 100% of notes assigned to the feature-absent note-level phenotype. Secondly, it includes the patients who have higher proportions of ‘GI notes’ than ‘classic notes’ from the PAM model. Patient ID 6 appears to be an exception, as it contains more ‘classic notes’ than ‘GI notes.’

The classic patient-level phenotype also captures several groups of patients. First, it includes patients with no notes assigned to the GI note-level phenotype. It also contains patients with higher or equal proportions of “classic notes” than “GI notes.” Patient ID 20 appears to be an exception, as it is the only individual in this patient-level phenotype without any “classic notes.”

## 3. Discussion

In this study, we characterize symptom phenotypes in adolescent endometriosis using two different approaches to clustering: note-level (PAM) and patient-level (MGM). We identify three phenotypes at the note-level (classic, GI, and feature-absent) and two phenotypes at the patient-level (classic and non-classic), based on the predictors of pelvic pain, chronic pain, dysmenorrhea, and GI symptoms. To our knowledge, this is the first study to compare different clustering techniques for phenotype evaluation in endometriosis.

Our findings indicate that both clustering methods identify a distinct classic phenotype, representing the traditional symptoms of endometriosis. In the note-level analysis, this phenotype is characterized by pelvic pain, chronic pain, and dysmenorrhea. At the patient-level, the corresponding phenotype is more specifically defined by chronic pain and pelvic pain, with dysmenorrhea present in both patient-level phenotypes but not predictive of either. These findings align with existing literature, which identifies pelvic pain as the most common presenting symptom of endometriosis^42^. For instance, prior studies have found nearly two-thirds of adolescents with chronic acyclic pelvic pain end up having laparoscopic evidence of endometriosis^43, 44^. Chronic pain is also a well-established factor, as prior work highlights the chronicity and severity of pain as a key driver for adolescents to seek medical care, particularly given the long diagnostic delays in this patient population^43^.

The contribution of dysmenorrhea to our classic phenotypes is particularly noteworthy. Prior literature identifies cyclic versus non-cyclic pain as a distinguishing factor between adolescent and adult endometriosis^42, 45^. While cyclic pain (dysmenorrhea) is a well-established presenting symptom in adult endometriosis, it is less prominent in adolescents. One study reported that only 9.4% of adolescents with endometriosis presented with cyclic pain alone, compared to 90.6% who had acyclic pain^45, 46^. Although this analysis does not directly compare adults and adolescents, the patient-level findings suggest that dysmenorrhea is not a major contributor to either phenotype, despite being a somewhat prevalent symptom (although less prevalent than acyclic pain and GI symptoms). Additionally, the patient-level classic phenotype is associated with higher therapy utilization (hormonal, IUP, or OCP) compared to the mixed phenotype, further supporting the notion that adolescents with more typical symptoms are more likely to receive targeted treatments.

Next, both clustering methods identify a phenotype with a higher contribution of GI symptoms. The note-level analysis uniquely identifies a distinct GI phenotype, while the patient-level analysis captures these symptoms within a broader non-classic phenotype, where GI symptoms are the largest contributing feature. This finding aligns with prior studies showing that adolescents with endometriosis can present with GI symptoms, although less commonly than pelvic pain^42, 46^. For example, a prior study found that nausea was reported more frequently by adolescents than adults ( 70% vs. 51%)^43^, underscoring the importance of recognizing GI symptom dominant presentations in the population.

Notably, for both clustering methods, the GI/non-classic phenotype is less likely to be associated with treatments (See **Supplementary Tables 6 and 7**), in line with prior evidence that adolescents with predominantly GI or non-classic presentations often experience delays in diagnosis. That is, previous studies reported that their symptoms are frequently attributed to functional bowel disorders or lead to GI specialty referrals, resulting in fragmented care and missed opportunities for early intervention^13 47^.

The note-level clustering approach identifies a third phenotype for whom none of the four indicator variables appear, hence we refer to it as “feature-absent”. These notes likely reflect periods of symptom quiescence, underreporting, or notes unrelated to endometriosis. Interestingly, the feature-absent phenotype is more likely to be associated with hormonal, OCP or IUD therapy, suggesting that at least part of this group reflects periods of symptom quiescence during treatment. This supports prior literature demonstrating that therapies, including OCPs, are often effective in relieving pain^48^. In our post-hoc investigation of this possibility, we repeated the analysis after removing notes where all 12 annotated features were absent. The results (not reported herein) with respect to phenotype descriptions and associations with therapy remained unchanged. This further alludes to the ability of this clustering approach to capture stable or asymptomatic periods. Additionally, the feature-absent comprised over 75% of the total notes in the study sample, consistent with prior report that adolescents with endometriosis are frequent overall users of healthcare^49^.

From a methodological standpoint, this study highlights the complementary perspectives the note-level and patient-level analyses can provide. The patient-level approach emphasizes between-patient variability, summarizing overall symptom profiles across all notes for a given patient. This method captures the well-established variability disease presentation among individuals^42, 45^. In contrast, the note-level approach captures the temporality and within-patient heterogeneity of symptoms, providing a dynamic view of symptom fluctuations over time. While evidence for temporal variability is more limited^14^, prior work suggests that changing symptom patterns may be linked to pathologic differences in endometriosis lesions between adolescents and adults or increased prevalence of deep infiltrating endometriosis as patients age^43^. Our findings quantify these within-patient fluctuations through note-level clustering, contributing to previous literature noting symptom heterogeneity across time. Together, these approaches highlight the value of capturing both episodic and longitudinal symptom patterns to improve phenotypic characterization.

A major strength of this study is its use of unstructured EHR data, capturing real-world symptom descriptions that are often overlooked when relying only on structured data and diagnostic codes. Second, the use of two different clustering approaches allows us to identify clinically meaningful phenotypes that capture both within-and between-patient variability, offering a more comprehensive symptom profile than a single method in isolation. Clinically, our findings highlight the importance of recognizing atypical presentations, such as GI-predominant symptoms and cases where the four indicator variables are absent, as these patients are more likely to face diagnostic delays or unnecessary referrals.

## 4. Limitations

Despite its strengths and novelty, our study has several limitations. First, the small sample size, while sufficient for a proof-of-concept analysis, limits the generalizability of our findings. Additionally, the present study focuses on symptom features without incorporating other clinical variables, such as imaging findings, genetic components, or treatment responses that are becoming increasingly studied. Repeating these analyses in larger samples, as well as conducting multi-site studies can enable more comprehensive comparisons of the identified phenotypes. Furthermore, future research can benefit from integrating additional clinical variables to identify more nuanced symptom patterns or potential moderators, as well as compare them to adult populations.

## 5. Conclusion

In conclusion, this study demonstrates the utility of unsupervised clustering methods to characterize symptom heterogeneity in adolescent endometriosis using EHR-derived clinical notes. Both clustering approaches identify a distinct classic phenotype with hallmark pain symptoms and a GI/non-classic phenotype, highlighting the diverse symptom presentations in this population. The note-level analysis uniquely identifies a feature-absent phenotype, reflecting periods of symptom dormancy, underreporting, or unrelated notes. By capturing both episodic within-patient variability and between-patient heterogeneity, our findings support the dynamic nature of adolescent endometriosis. These results emphasize the importance of recognizing atypical presentations, which will ultimately aid in earlier diagnosis.

## Data Availability

All data produced in the present study are not publicly available due to patient privacy, security, and the Health Insurance Portability and Accountability Act of 1996 (HIPAA) requirement.

## Acknowledgements

This work was supported in part through the use of research platform AI-Ready Mount Sinai (AIR.MS) and the expertise provided by the team at the Hasso Plattner Institute for Digital Health at Mount Sinai (HPI.MS), as well as the Office of Research Infrastructure of the National Institutes of Health under award number S10OD026880. This work was also supported in part through the computational and data resources and staff expertise provided by Scientific Computing and Data at the Icahn School of Medicine at Mount Sinai and supported by the Clinical and Translational Science Award (CTSA) grant UL1TR004419 from the National Center for Advancing Translational Sciences.

## References

1. ACOG Committee Opinion No. 760: Dysmenorrhea and Endometriosis in the Adolescent. Obstet Gynecol. 2018;132(6):e249-e258.

2. Khashchenko EP, Uvarova EV, Fatkhudinov TK, et al. Endometriosis in Adolescents: Diagnostics, Clinical and Laparoscopic Features. J Clin Med. 2023;12(4).

3. Dun EC, Kho KA, Morozov VV, et al. Endometriosis in adolescents. Jsls. 2015;19(2).

4. Balun J, Dominick K, Cabral MD, et al. Endometriosis in adolescents: a narrative review. Pediatric Medicine. 2019;2.

5. Nielsen LJ, Poulsen K, Funch AL, et al. The lived experiences of endometriosis in adolescence—A critical hermeneutic perspective. Scandinavian Journal of Caring Sciences. 2023;37(4):1038–1047.

6. Nakamura T. Clinical Aspects of Adolescent Endometriosis. Endocrines. 2021;2(3):301–310.

7. Zhan W, Wu F, Zhang Y, et al. Identification of cough-variant asthma phenotypes based on clinical and pathophysiologic data. J Allergy Clin Immunol. 2023;152(3):622–632.

8. Moore WC, Meyers DA, Wenzel SE, et al. Identification of asthma phenotypes using cluster analysis in the Severe Asthma Research Program. Am J Respir Crit Care Med. 2010;181(4):315–323.

9. Takeshita S, Nishioka Y, Tamaki Y, et al. Novel subgroups of obesity and their association with outcomes: a data-driven cluster analysis. BMC Public Health. 2024;24(1):124.

10. Ocay DD, Loewen A, Premachandran S, et al. Psychosocial and psychophysical assessment in paediatric patients and young adults with chronic back pain: A cluster analysis. Eur J Pain. 2022;26(4):855–872.

11. Alter BJ, Anderson NP, Gillman AG, et al. Hierarchical clustering by patient-reported pain distribution alone identifies distinct chronic pain subgroups differing by pain intensity, quality, and clinical outcomes. PLoS One. 2021;16(8):e0254862.

12. Ensari I, Caceres BA, Jackman KB, et al. Digital phenotyping of sleep patterns among heterogenous samples of Latinx adults using unsupervised learning. Sleep Med. 2021;85:211–220.

13. de Sanctis V, Matalliotakis M, Soliman AT, et al. A focus on the distinctions and current evidence of endometriosis in adolescents. Best Practice & Research Clinical Obstetrics & Gynaecology. 2018;51:138–150.

14. Ensari I, Pichon A, Lipsky-Gorman S, et al. Augmenting the Clinical Data Sources for Enigmatic Diseases: A Cross-Sectional Study of Self-Tracking Data and Clinical Documentation in Endometriosis. Appl Clin Inform. 2020;11(5):769–784.

15. Bittelbrunn CC, de Fraga R, Martins C, et al. Pelvic floor physical therapy and mindfulness: approaches for chronic pelvic pain in women—a systematic review and meta-analysis. Archives of Gynecology and Obstetrics. 2023;307(3):663–672.

16. Horne EMF, McLean S, Alsallakh MA, et al. Defining clinical subtypes of adult asthma using electronic health records: Analysis of a large UK primary care database with external validation. International Journal of Medical Informatics. 2023;170:104942.

17. Florensa D, Mateo-Fornés J, Solsona F, et al. Use of Multiple Correspondence Analysis and K-means to Explore Associations Between Risk Factors and Likelihood of Colorectal Cancer: Cross-sectional Study. J Med Internet Res. 2022;24(7):e29056.

18. Maugis C, Celeux G, Martin-Magniette M-L. Variable Selection for Clustering with Gaussian Mixture Models. Biometrics. 2009;65(3):701–709.

19. Abdi H, Valentin D. Multiple correspondence analysis. Encyclopedia of measurement and statistics. 2007;2(4):651–657.

20. Rodriguez-Sabate C, Morales I, Sanchez A, et al. The Multiple Correspondence Analysis Method and Brain Functional Connectivity: Its Application to the Study of the Non-linear Relationships of Motor Cortex and Basal Ganglia. Front Neurosci. 2017;11:345.

21. Sourial N, Wolfson C, Zhu B, et al. Correspondence analysis is a useful tool to uncover the relationships among categorical variables. J Clin Epidemiol. 2010;63(6):638–646.

22. Higgs NT. Practical and Innovative Uses of Correspondence Analysis. Journal of the Royal Statistical Society: Series D (The Statistician). 1991;40(2):183–194.

23. Partitioning Around Medoids (Program PAM). Finding Groups in Data 1990:68-125.

24. Oehm AW, Springer A, Jordan D, et al. A machine learning approach using partitioning around medoids clustering and random forest classification to model groups of farms in regard to production parameters and bulk tank milk antibody status of two major internal parasites in dairy cows. PLoS One. 2022;17(7):e0271413.

25. Gu Y, Preisser JS, Zeng D, et al. PARTITIONING AROUND MEDOIDS CLUSTERING AND RANDOM FOREST CLASSIFICATION FOR GIS-INFORMED IMPUTATION OF FLUORIDE CONCENTRATION DATA. Ann Appl Stat. 2022;16(1):551–572.

26. Maechler M, Rousseeuw P, Struyf A, et al. cluster: Cluster analysis basics and extensions (R package version 2.1.1). 2021.

27. Schubert E, Rousseeuw PJ. Faster k-Medoids Clustering: Improving the PAM, CLARA, and CLARANS Algorithms. Cham: Springer International Publishing 2019:171-187.

28. Mushtaq H, Khawaja SG, Akram MU, et al. A Parallel Architecture for the Partitioning Around Medoids (PAM) Algorithm for Scalable Multi-Core Processor Implementation with Applications in Healthcare. Sensors (Basel). 2018;18(12).

29. Bhandari N, Pahwa P. Evaluating Partitioning Based Clustering Methods for Extended Non-negative Matrix Factorization (NMF). Intelligent Automation & Soft Computing. 2023;35(2).

30. Leis AM, McSpadden E, Segaloff HE, et al. K-medoids clustering of hospital admission characteristics to classify severity of influenza virus infection. Influenza and Other Respiratory Viruses. 2023;17(3):e13120.

31. Maechler M. Cluster analysis extended Rousseeuw et al. R CRAN. 2013.

32. Rousseeuw PJ. Silhouettes: A graphical aid to the interpretation and validation of cluster analysis. Journal of Computational and Applied Mathematics. 1987;20:53–65.

33. Kaufman L, Rousseeuw PJ. Finding groups in data: an introduction to cluster analysis: John Wiley & Sons 2009.

34. Lun A. Bootstrapping for cluster stability Available: https://ltla.github.io/SingleCellThoughts/general/bootstrapping.html.

35. Komárek A. A new R package for Bayesian estimation of multivariate normal mixtures allowing for selection of the number of components and interval-censored data. Computational Statistics & Data Analysis. 2009;53(12):3932–3947.

36. Komárek A, Komárková L. Clustering for multivariate continuous and discrete longitudinal data. The Annals of Applied Statistics. 2013;7(1):177–200, 124.

37. Link WA, Eaton MJ. On thinning of chains in MCMC. Methods in Ecology and Evolution. 2012;3(1):112–115.

38. Aitkin M, Liu CC, Chadwick T. Bayesian model comparison and model averaging for small-area estimation. The Annals of Applied Statistics. 2009;3(1):199–221, 123.

39. Plummer M. Penalized loss functions for Bayesian model comparison. Biostatistics. 2008;9(3):523–539.

40. Vehtari A, Gelman A, Simpson D, et al. Rank-normalization, folding, and localization: An improved R for assessing convergence of MCMC (with discussion). Bayesian analysis. 2021;16(2):667–718.

41. Gelman A, Carlin JB, Stern HS, et al. Bayesian data analysis: Chapman and Hall/CRC 1995.

42. Laufer MR, Sanfilippo J, Rose G. Adolescent Endometriosis: Diagnosis and Treatment Approaches. Journal of Pediatric and Adolescent Gynecology. 2003;16(3, Supplement):S3–S11.

43. DiVasta AD, Vitonis AF, Laufer MR, et al. Spectrum of symptoms in women diagnosed with endometriosis during adolescence vs adulthood. American Journal of Obstetrics and Gynecology. 2018;218(3):324.e321-324.e311.

44. Janssen EB, Rijkers ACM, Hoppenbrouwers K, et al. Prevalence of endometriosis diagnosed by laparoscopy in adolescents with dysmenorrhea or chronic pelvic pain: a systematic review. Human Reproduction Update. 2013;19(5):570–582.

45. Shim JY, Laufer MR. Adolescent Endometriosis: An Update. Journal of Pediatric and Adolescent Gynecology. 2020;33(2):112–119.

46. Laufer MR, Goitein L, Bush M, et al. Prevalence of Endometriosis in Adolescent Girls With Chronic Pelvic Pain Not Responding to Conventional Therapy. Journal of Pediatric and Adolescent Gynecology. 1997;10(4):199–202.

47. MAROUN P, COOPER MJW, REID GD, et al. Relevance of gastrointestinal symptoms in endometriosis. Australian and New Zealand Journal of Obstetrics and Gynaecology. 2009;49(4):411–414.

48. Ballard K, Lowton K, Wright J. What’s the delay? A qualitative study of women’s experiences of reaching a diagnosis of endometriosis. Fertility and Sterility. 2006;86(5):1296–1301.

49. Eisenberg VH, Decter DH, Chodick G, et al. Burden of Endometriosis: Infertility, Comorbidities, and Healthcare Resource Utilization. J Clin Med. 2022;11(4).

