## Supplementary Materials for "Phenotyping Adolescent Endometriosis: Characterizing Symptom Heterogeneity Through Note- and Patient-Level Clustering"

**Adolescent Endometriosis Clustering**

**Supplementary Material 1 – Study Data and Methods**

**Supplementary Table 1.** The number of notes (frequency) with a mention of each feature and the associated percent frequency. Bolded values indicate a percent frequency < 10%.

| **Feature** | **Frequency (N)** | **Percent frequency (%)** |
| --- | --- | --- |
| Endometriosis | 66 | 11.62 |
| Pelvic pain | 94 | 16.55 |
| Pelvic tenderness | 33 | **5.81** |
| Abdominal pain | 120 | 21.13 |
| Abdominal tenderness | 35 | **6.16** |
| Dysmenorrhea | 84 | 14.79 |
| Dyspareunia | 11 | **1.94** |
| Gastrointestinal symptoms | 125 | 22.01 |

**Supplementary Table 2.** Features associated with the selected dimension including their coordinates (i.e., correlations) within each dimension, their total contributions, and the quality of the total contributions (i.e., cosine 2).

| Predictor | Category | Dim 1 | Dim 2 | Dim 3 | Dim 4 | Contribution | Cosine 2 |
| --- | --- | --- | --- | --- | --- | --- | --- |
| Endometriosis | 0 | -0.19 | 0.07 | 0.2 | -0.02 | 5.32 | 0.75 |
| Endometriosis | 1 | 1.77 | -0.69 | -1.87 | 0.19 | 50.79 | 0.75 |
| Pelvic pain | 0 | -0.29 | 0.07 | 0.03 | -0.14 | 5.42 | 0.71 |
| Pelvic pain | 1 | 1.87 | -0.42 | -0.19 | 0.87 | 34.69 | 0.71 |
| Abdominal pain | 0 | -0.21 | -0.33 | 0.02 | -0.04 | 8.73 | 0.76 |
| Abdominal pain | 1 | 1.02 | 1.59 | -0.09 | 0.19 | 41.86 | 0.76 |
| GI symptoms | 0 | -0.2 | -0.34 | 0.01 | 0.11 | 10.08 | 0.78 |
| GI symptoms | 1 | 0.93 | 1.57 | -0.03 | -0.49 | 45.96 | 0.78 |
| Dysmenorrhea | 0 | -0.22 | 0.11 | -0.08 | 0.25 | 10.71 | 0.95 |
| Dysmenorrhea | 1 | 1.57 | -0.81 | 0.6 | -1.85 | 77.95 | 0.95 |
| Severe pain | 0 | -0.11 | 0.02 | -0.15 | -0.07 | 3.44 | 0.86 |
| Severe pain | 1 | 2.31 | -0.37 | 3.32 | 1.4 | 73.66 | 0.86 |
| Chronic pain | 0 | -0.26 | 0.08 | 0.02 | -0.01 | 2.85 | 0.74 |
| Chronic pain | 1 | 2.59 | -0.76 | -0.23 | 0.07 | 28.52 | 0.74 |

GI: gastrointestinal

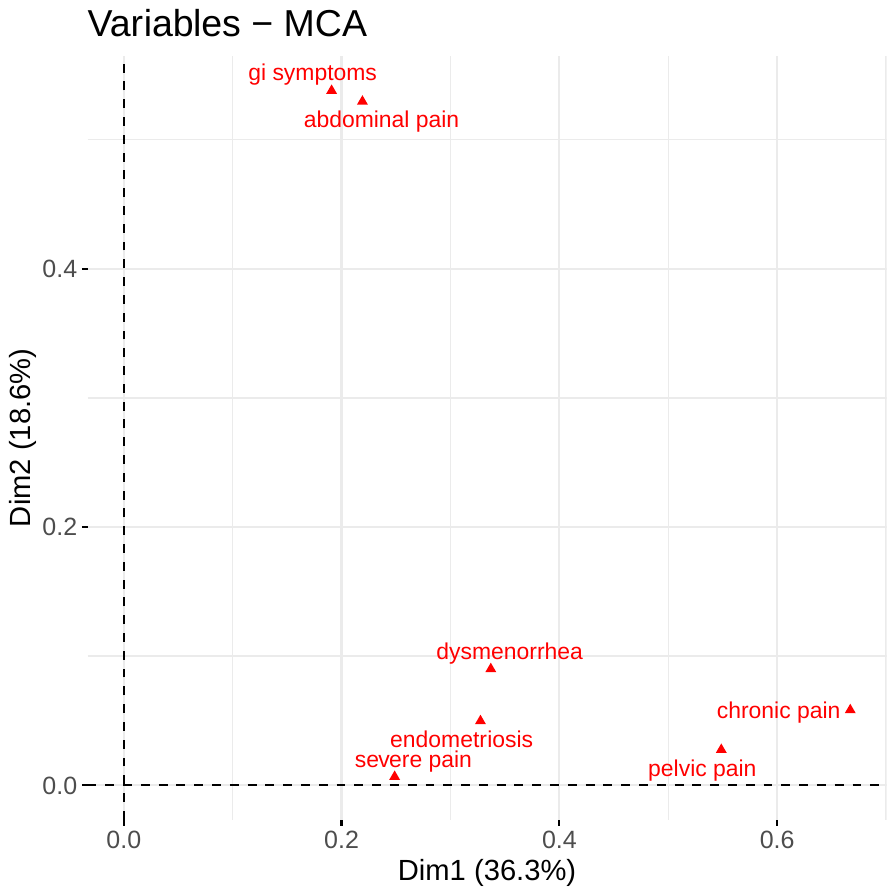

**Supplementary Figure 1.** Correspondence analysis map in the first 2 dimensions. GI: gastrointestinal

**Supplementary Table 3.** Pearson's rho correlation coefficients for the multivariate correlation

|  | Pelvic pain | Abdominal pain | Chronic pain | GI symptoms | dysmenorrhea | Severe pain | endometriosis |
| --- | --- | --- | --- | --- | --- | --- | --- |
| Pelvic pain | 1 |  |  |  |  |  |  |
| Abdominal pain | 0.25*** | 1 |  |  |  |  |  |
| Chronic pain | 0.56*** | 0.2*** | 1 |  |  |  |  |
| GI symptoms | 0.12* | 0.49*** | 0.18*** | 1 |  |  |  |
| dysmenorrhea | 0.29*** | 0.06 | 0.44*** | 0.11* | 1 |  |  |
| Severe pain | 0.28*** | 0.12* | 0.32*** | 0.13** | 0.24*** | 1 |  |
| endometriosis | 0.35*** | 0.1** | 0.44*** | 0.13** | 0.21*** | 0.07 | 1 |

*p<0.05, **p<0.001, ***p<0.0001

GI: gastrointestinal

**Supplementary Material 2. Model Results**

**Supplementary Table 4.** Metrics for PAM clustering for different numbers of clusters (k)

| Metric | K=2 | K=3 | K=4 |
| --- | --- | --- | --- |
| Point variability | 74.3 % | 74.3% | NA |
| Avg. Silhouette width | 0.67 | 0.76 | 0.79 |
| WCSS | 0.27 | 0.19 | 0.15 |
| Jaccard Index avg  (min, max) | 0.70  (0.57, 0.83) | 0.68  (0.53, 0.89) | 0.80  (0.39, 1.00) |

WCSS: within-cluster sum of squares

**Supplementary Table 5**. Note-level clustering model evaluation for the best-fitting model with K = 3.

| Metric | Feature absent phenotype | Classic phenotype | GI phenotype | Average/overall |
| --- | --- | --- | --- | --- |
| Point variability | NA | NA | NA | 74.3% |
| Silhouette width | 0.84 | 0.39 | 0.51 | 0.76 |
| WCSS | NA | NA | NA | 0.19 |
| Jaccard Index | 0.89 | 0.53 | 0.63 | 0.68 |

WCSS: within-cluster sum of squares

**Supplementary Table 6.** Results of the chi-squared analysis of annotated across the three note-level phenotypes.

| Phenotypes | Feature-Absent  N=530 notes | | | Classic  N = 57 Notes | | GI Symptoms  N = 108 Notes | | |
| --- | --- | --- | --- | --- | --- | --- | --- | --- |
|  | % Freq | | *X*^2^ statistic | % Freq | *X*^2^ statistic | % Freq | | *X*^2^ statistic |
| Pelvic pain | 5.3 | -1.165 | | 89.5 | 5.387*** | 13.9 | -3.984** | |
| Dysmenorrhea | 4.9 | -0.870 | | 70.2 | 3.713 | 16.7 | -2.680 | |
| Chronic pain | 0.4 | -5.441*** | | 93.0 | 9.659*** | 7.41 | -3.785 | |
| GI symptoms | 0.0 | -8.570*** | | 29.8 | -4.051** | 100.0 | 12.462*** | |
| Endometriosis | 4.2 | -0.355 | | 50.9 | 2.608 | 13.9 | -2.138 | |
| Pelvic tenderness | 2.8 | 1.221 | | 14.0 | -0.675 | 9.26 | -0.578 | |
| Abdominal pain | 8.5 | 0.502 | | 31.6 | -3.615 | 52.8 | 2.955 | |
| Abdominal tenderness | 2.64 | 0.577 | | 8.8 | -1.999 | 14.8 | 1.334 | |
| Dyspareunia | 0.4 | -1.197 | | 8.8 | 1.159 | 3.7 | 0.091 | |
| Severe pain | 1.7 | -0.746 | | 28.1 | 2.714 | 5.6 | -1.848 | |
| Pain medication |  |  | |  |  |  |  | |
| Over the counter | 3.0 | -0.405 | | 38.6 | 2.385 | 10.2 | -1.875 | |
| Prescription | 7.4 | 2.862 | | 43.9 | 0.571 | 12.0 | -3.415* | |
| Any pain med | 9.1 | 2.537 | | 68.4 | 1.976 | 13.9 | -4.432** | |
| Therapy |  |  | |  |  |  |  | |
| OCP | 13.4 | 4.927*** | | 54.4 | -1.373 | 24.1 | -3.625 | |
| IUD | 5.3 | 0.542 | | 28.1 | -1.460 | 26.9 | 0.853 | |
| Hormonal therapy | 9.4 | 6.704*** | | 15.8 | -3.006 | 8.3 | -3.843** | |
| Any therapy | 25.9 | 7.999*** | | 77.2 | -3.822** | 49.2 | -4.361** | |

*p<0.05, **p<0.01, ***p<0.001

GI: gastrointestinal; IUD: intrauterine device; OCP: oral contraceptive pill

**Supplementary Table 7.** Results of the independent t-tests comparing mean feature proportions between the phenotypes identified via patient-level model. Statistically significant differences are bolded.

| Characteristic | Non-Classic Phenotype  (N=13 patients)^1^ | Classic Phenotype  (N=13 patients) | t-statistic | p-value |
| --- | --- | --- | --- | --- |
| Pelvic pain | 0.03 ± 0.04 | 0.33 ± 0.12 | 8.369 | **<0.001** |
| Dysmenorrhea | 0.13 ± 0.19 | 0.23 ± 0.18 | 1.387 | 0.178 |
| Chronic pain | 0.03 ± 0.05 | 0.24 ± 0.17 | 4.398 | **0.001** |
| GI symptoms | 0.14 ± 0.23 | 0.18 ± 0.17 | 0.522 | 0.607 |
| Endometriosis | 0.07 ± 0.10 | 0.26 ± 0.23 | 2.727 | **0.015** |
| Pelvic tenderness | 0.00 ± 0.01 | 0.08 ± 0.11 | 2.372 | **0.035** |
| Abdominal pain | 0.05 ± 0.10 | 0.10 ± 0.12 | 1.139 | 0.266 |
| Abdominal tenderness | 0.01 ± 0.02 | 0.04 ± 0.06 | 1.656 | 0.119 |
| Dyspareunia | 0.00 ± 0.01 | 0.06 ± 0.12 | 1.566 | 0.143 |
| Severe pain | 0.05 ± 0.06 | 0.09 ± 0.11 | 1.006 | 0.328 |
| Pain medication | 0.03 ± 0.04 | 0.16 ± 0.24 | 1.951 | 0.074 |
| Over the counter | 0.07 ± 0.12 | 0.21 ± 0.24 | 1.811 | 0.088 |
| Prescription | 0.09 ± 0.12 | 0.27 ± 0.25 | 2.311 | **0.034** |
| Any pain med | 0.27 ± 0.31 | 0.30 ± 0.22 | 0.274 | 0.787 |
| Therapy | 0.08 ± 0.14 | 0.17 ± 0.26 | 1.214 | 0.240 |
| OCP | 0.14 ± 0.25 | 0.10 ± 0.16 | -0.555 | 0.585 |
| IUD | 0.42 ± 0.29 | 0.46 ± 0.28 | 0.354 | 0.726 |
| Hormonal therapy | 0.07 ± 0.10 | 0.26 ± 0.23 | 2.727 | **0.015** |
| Any therapy | 0.00 ± 0.01 | 0.08 ± 0.11 | 2.372 | **0.035** |

^1^mean proportion of mentions per patient ± standard deviation

GI: gastrointestinal; IUD: intrauterine device; OCP: oral contraceptive pill
